# The end of the social confinement in Spain and the COVID-19 re-emergence risk

**DOI:** 10.1101/2020.04.14.20064766

**Authors:** Leonardo López, Xavier Rodó

## Abstract

After the spread of SARS-CoV-2 epidemic out of China, the world approaches the 2 million declared infected cases and death toll rises well above the 100 thousand. The course of pandemic evolution has shown great differences among countries and not much is yet known about the level of generated immunity, which might appear not to be long-lasting. In this situation, management of a recurrent disease seems to be a plausible scenario that countries worldwide will have to face, before effective drugs or a vaccine appear. Spain in Europe, appears to be the first country deciding to partly lift the strict social distancing regulations imposed. Whether this action may lead to further epidemic recrudescence, to a following second wave of cases or conversely, help return to previous normality, is a subject of great debate and interest to all other countries affected by COVID-19. Here we applied a modified *SEIR* compartmental model accounting for the spread of infection during the latent period, in which we had also incorporated effects of social confinement. We now modify this previous model configuration to mimic potential post-confinement scenarios, by simulating from instant massive liberation of different portions of the confined population, up to a more gradual incorporation of people to work. Results show how current lockdown conditions should be extended at least two weeks more to prevent a new escalation in cases and deaths, as well as a larger second wave occurring in just a few months. Conversely, best-case scenario in terms of lower COVID-19 incidence and casualties should gradually incorporate workers back in a daily proportion at most 30 percent higher than that of previous confinement. The former should begin not earlier than by the end of April and it would represent approximately 600 thousand people or a 3.75% rate for the whole of Spain.

## Introduction

COVID-19 pandemic has entered a new stage in which the United States appears to be at the epicenter and Europe seems to have passed the peak of SARS-CoV-2 outbreak^1^. In this situation and whereas Italy decided not to yet lift lockdown measures, Spain after a large outbreak of still tens of thousands active cases, already plans for immediate DE confinement actions. Data on the last similar scale pandemic more than one century ago remains limited, despite aftermath reports exist^2^. The fact that the 1918 Spanish flu pandemic had a second deadly epidemic wave, presumably caused by strain mutations^3^, has also stimulated a vivid debate on whether actions to take now should or not incorporate this uncertain future. Under this scenario, optimal interventions on the COVID-19 pandemic aiming at a relaxation of strict social distancing enforcement are hard to fathom. More so, when the extent of pre-symptomatic and asymptomatic infections is not narrowly constrained (e.g. may be up to 86 percent^4^). Europe’s worst hit countries, Italy and Spain, are now seeing a flattening of their respective death tolls. On March 29, the Government of Spain published a decree that paralyzed all sectors considered non-essential in a draconian lockdown^5,6^. The objective was to take advantage of Easter holidays to transfer the low level of mobility from Saturdays and Sundays to the rest of the days of the week, thereby reducing new infections and thus giving hospitals and ICUs a break when on the brink of collapse. Under an impending economic crisis, the Spanish executive government now believes it is time for the economy to recover certain dynamism^7^. However, whether soon or later the return to normality should be attempted, the decision of when and how best decide to optimally move in that direction is at the center of worldwide debate^8^. This decision unfortunately presents a double problem in which two factors are related. First, there is a risk that the return to activity will end up causing a spike in infections that will aggravate the sanitary collapse at a time when the situation is still delicate. Second, there is no way to assess this risk, given the lack of existing information on the actual number of people infected or the real extent of immunity developed among the population. In both Spain and Italy active infections are still in the thousands, and a lift of lockdown makes the risk of a flare up in cases again a serious threat. According to the WHO, it is premature for Europe to think about a lack of confinement, given the number of active cases and waiting longer would be more prudent^9^. In the UK, for example, much of the industry is at a stop and these measures are expected to continue for at least two months^10^. On the other hand, confinement in the current circumstances seems to be working satisfactorily considering that the number of new infections and fatal cases is decreasing in these two countries. Re-allowing the mobility of a large number of the confined population might increase the risk that these figures will rise again. To this end, in this article, we present projections based on a modified SEIR model that allows simulating both the degree of population confinement and the different post-confinement scenarios. Our study intends to evaluate effects of different actionable scenarios for Spain and inform also other countries of the risk that each of these scenarios entails.

### Brief model description

We used a generalized *SEIR* modelling framework similar to Peng et al.^11^, that enables testing control interventions and that was recently implemented for Spain and Italy.^12^. A return function with *τ* from *C* (confined) to *S* (susceptible) was now added, through which the effect of population de-confinement with different scenarios can be explored. The general description of the model can be seen below,

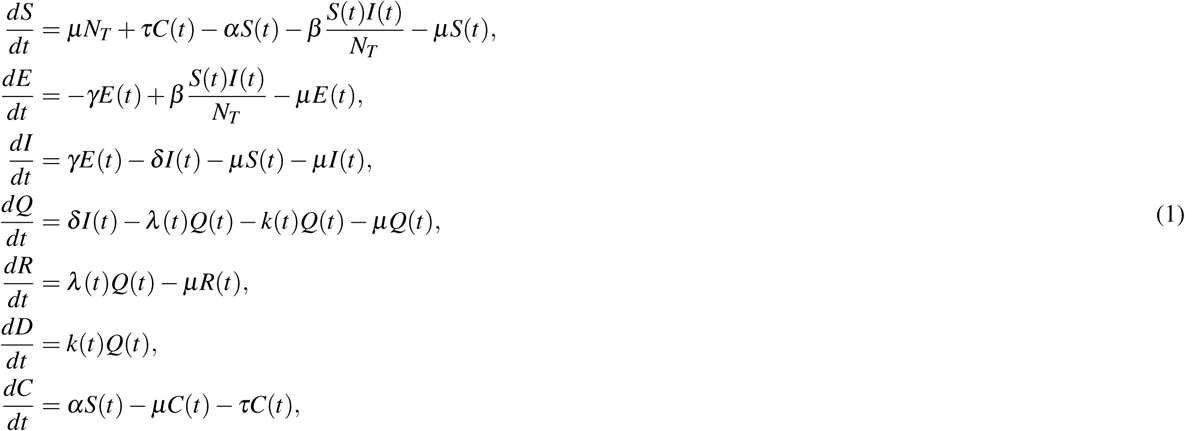

where,*S*(*t*) is the susceptible population, *C*(*t*) is the confined susceptible population, *E*(*t*) is the exposed population, *I*(*t*) is the infectious population, *Q*(*t*) is the population under Quarantine (infected reported cases), *R*(*t*) is the recovered population and *D*(*t*) is the dead population by the virus.

The main parameters of the model are the protection rate (*α*), the infection rate (*β*), the incubation rate (*γ*), the quarantine rate (*δ*), the natural death and birth rates (*µ*) (1*/*(80 365)), the recovery rate (*λ* (*t*)), the mortality rate by the virus (*k*(*t*)) and finally *τ* is the de-confinement rate. The parameters of the model are listed in table 1.

**Table 1.** Model fitted parameters

| $\alpha$ | $\beta$ | $\gamma$ | $\delta$ | $\lambda_0$ | $\lambda_1$ | $k_0$ | $k_1$ |
| --- | --- | --- | --- | --- | --- | --- | --- |
| 0,025 | 1,288 | 0,870 | 0,585 | 0,057 | 0,026 | 0,070 | 0,144 |

## Results

We simulated two different possible de-confinement strategies, namely the massive return of people to work on a given day and the gradual liberation of people from their confinement over a longer time interval. In the first strategy, the first case (Figure 1) is the sudden de-confinement of 30% of the population back to work, therefore these people is immediately integrated into the susceptible population. The simulation is generated for 30, 45 and 60 days after the confinement began on March 13, corresponding to April 13, 29 and May 14, respectively. The second case ((Figure 2)) is the sudden de-confinement of 40% of the protected population. As before, three different temporal scenarios (30, 45 and 60 days after the confinement of March 13 began) were again simulated for Spain, where 40% of all protected population is released. In the second strategy, instead, a gradual de-confinement rate is applied (Figure 3). The daily de-confinement rate in the first case studied is set to be 50% higher than the confinement rate. Finally, a daily rate 30% higher than the confinement rate was also simulated (Figure 4). The same three temporal scenarios are shown in these two latter situations (30, 45 and 60 days after the confinement began on March 13), but each of them is applied with the stated gradual de-confinement rates above.

**Figure 1.**
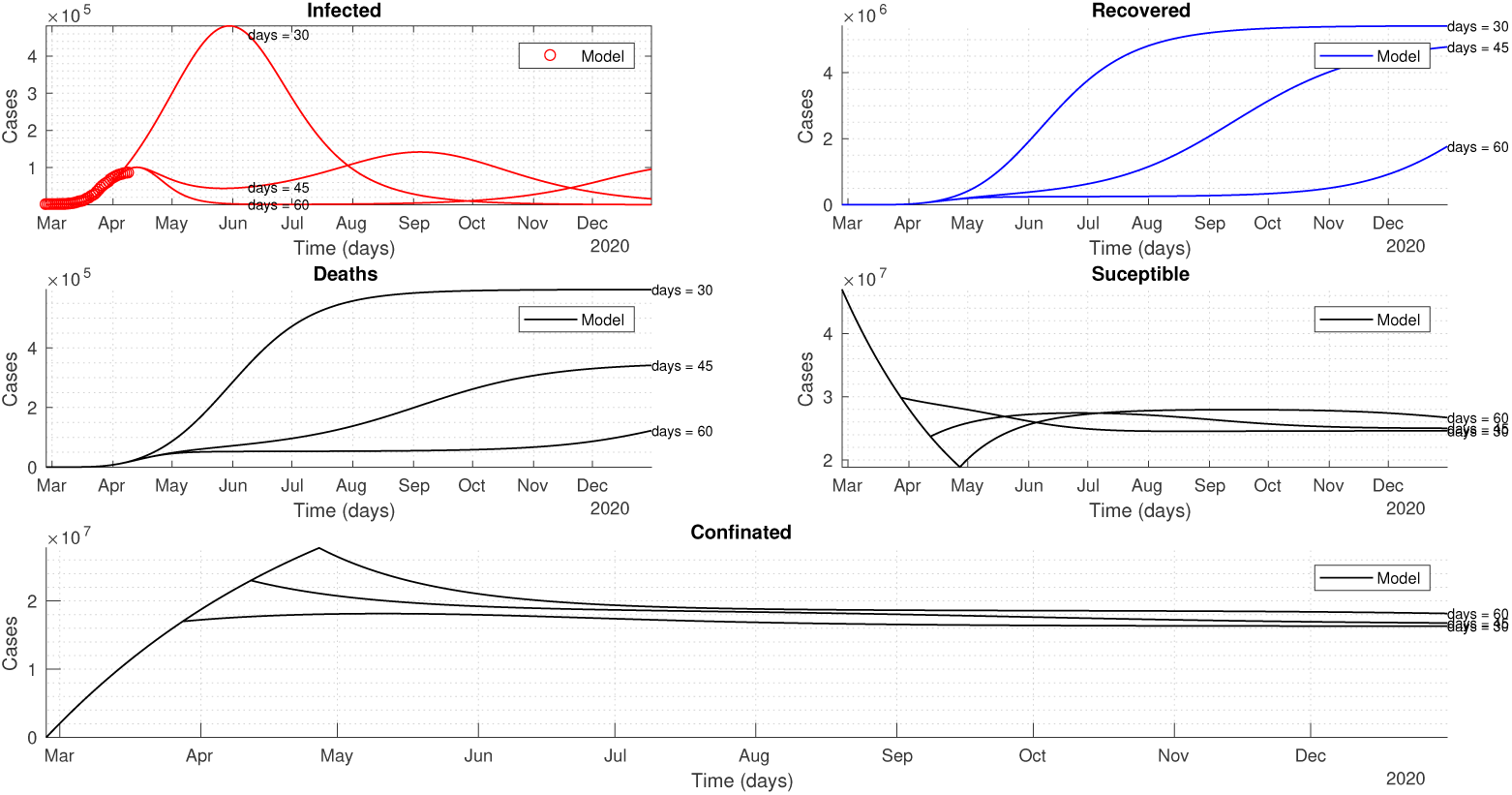
Sudden de-confinement of 30% of the population. Temporal dynamics is shown for the active cases, recovered population, deaths, susceptible and protected after 30, 45 and 60 days of confinement.

**Figure 2.**
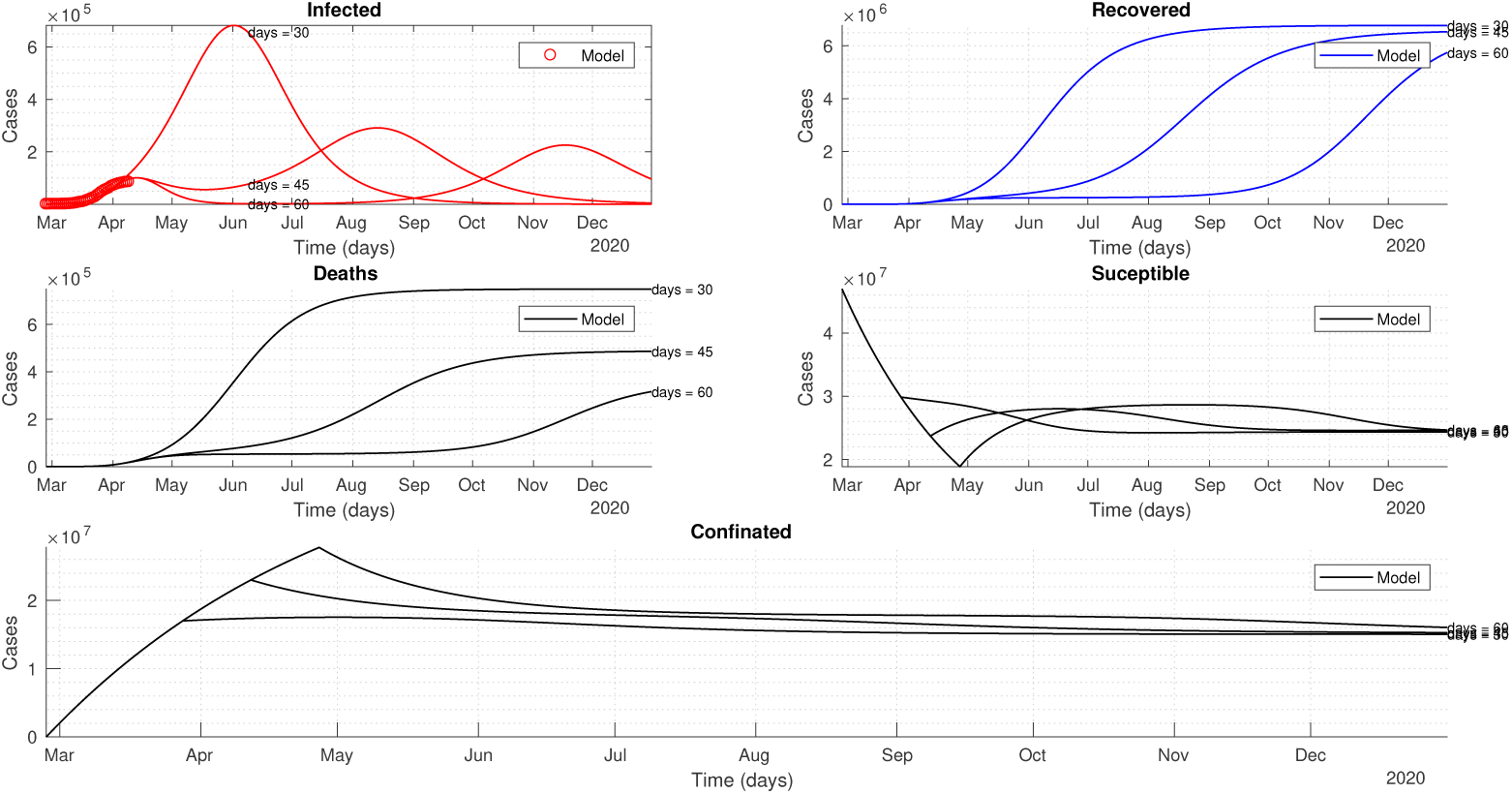
Sudden de-confinement of 40% of the population. Idem as for Figure 1.

**Figure 3.**
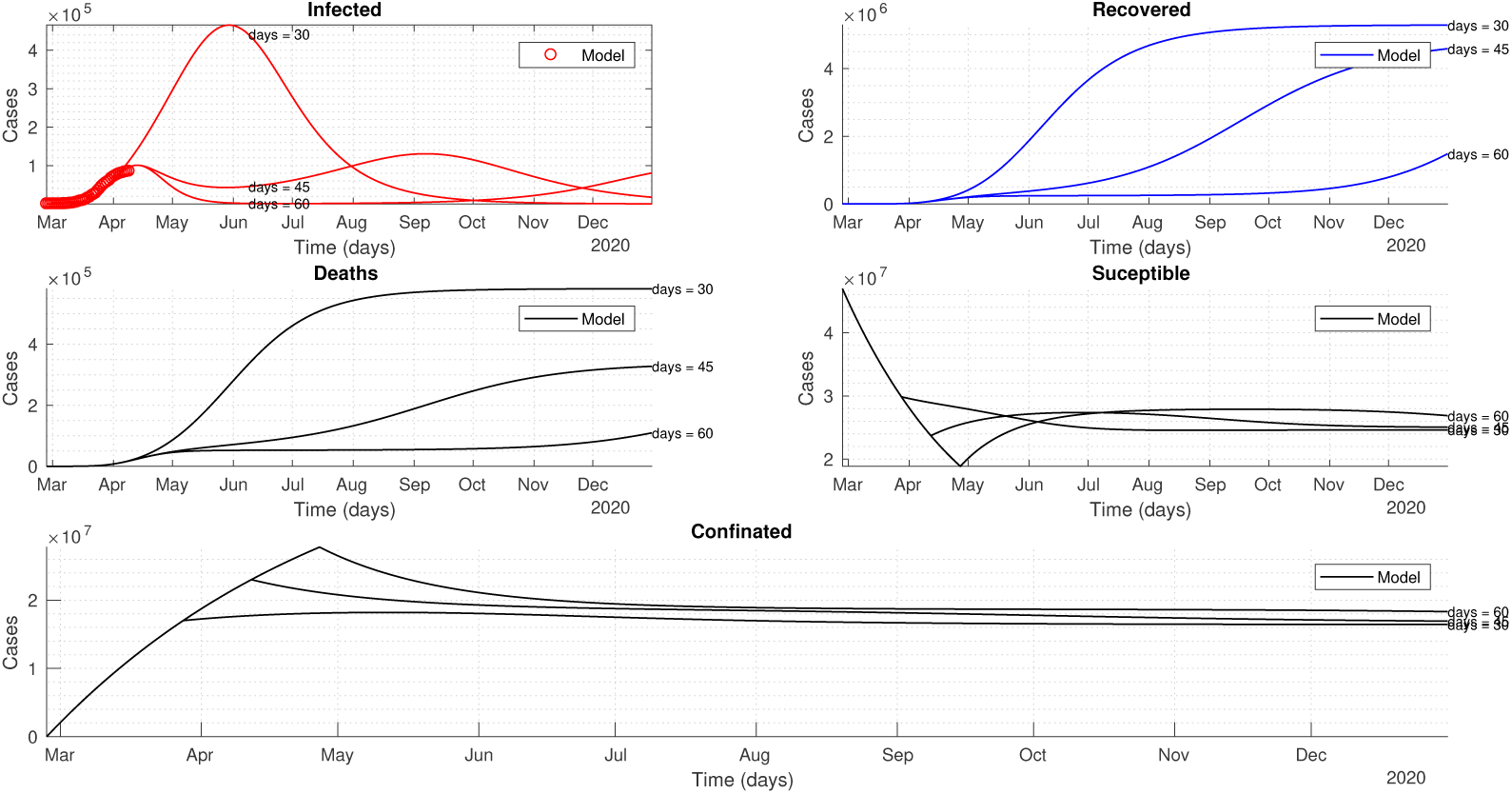
Gradual de-confinement with a rate 50% higher than the confinement rate. Temporal dynamics is shown for the active cases, recovered population, deaths, susceptible and protected after 30, 45 and 60 days of confinement.

**Figure 4.**
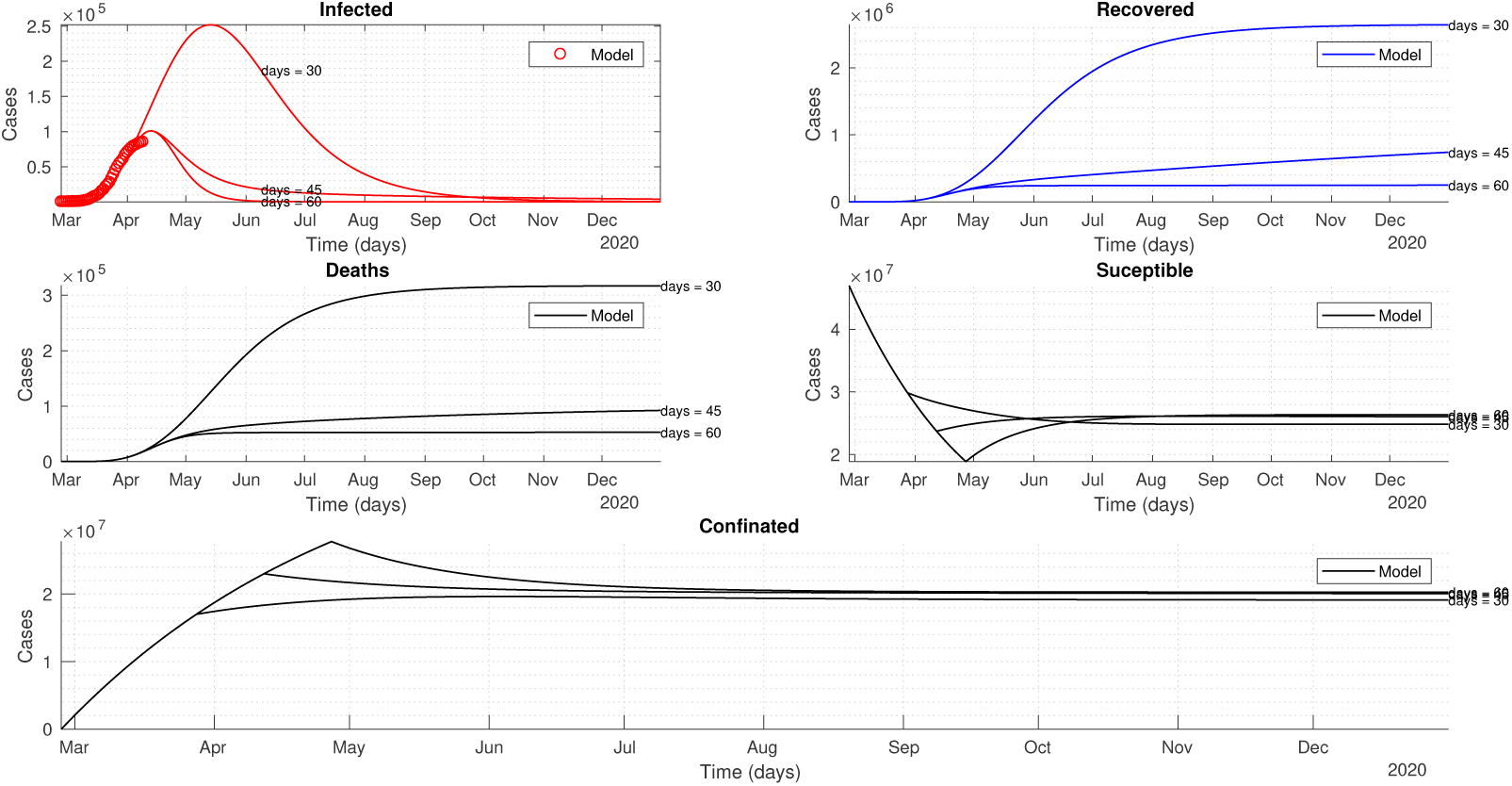
Gradual de-confinement with a rate 30% higher than the confinement rate. Dynamics of the active cases, recovered population, deaths, susceptible and protected after 30, 45 and 60 days of confinement.

## Discussion

The number of Infected represented in the previous figures corresponds to the number of active cases (and not new cases), understanding as such those people who are diagnosed with Covid-19 and who have not yet resolved the disease and, consequently, did not pass into the compartment of recovered population (*R*) or died (*D*) (it is for this reason that the peak in *I* occurs after that of new cases). In all cases, the release from confinement is applied on the days indicated in the figures, counted from March 13, so that 30 days correspond approximately to April 14, 45 days to April 29 and 60 days to May 14.

In the case of the sudden de-confinement of 30% of the population showed in Figure 1, it is observed how this involves the uninterrupted growth of the current wave and its continuity and amplification until the end of May. This means exceeding 400, 000 active cases at that date for all of Spain. On the contrary, the model shows how the maintenance of the confinement for 15 more days (April 29), involves the drastic limitation of the current epidemic peak to around 100, 000 active cases, as well as the generation of a second wave of cases peaking around mid-September. It should be noted that this second peak is delayed until 2021 when total confinement is extended to 60 days (May 14). A reduction in the number of deaths between the 30-day and 60-day scenarios to approximately half can be also seen in the central panel.

In figure 2, the most notable results are the growth in the final number of infected (from 400, 000 to 650, 000), unlike in the previous case. Of relevance also, is the appearance of a more prominent second peak in terms of active cases, that would occur prior to the scenario detailed above in figure 1. Under this assumption and even with a 60-day confinement, the second peak would occur in late 2020. Similarly, an increase in the number of deaths produced by Covid-19 is to be expected also.

Figure 3 shows important differences with regard to the previous scenarios where the de-confinement occurs suddenly. And most notably, even when compared to those that would be obtained with much longer total confinements. For example, at this gradual rate of de-confinement, started on day 30 from the start of confinement (March 13), the magnitude of the epidemic peak attained is comparable to that of a sudden de-confinement of around 40% of the confined population. This occurred 45 days after the start, on April 29. In contrast, the second peak in this progressive de-confinement at a rate of 50% higher than alpha since May 4, would reach a lower maximum number of active cases (110, 000 compared to 250, 000) than with the sudden release of 40% of the population confined on April 29. The total change in the number of deaths is also substantial between this scenario and the previous ones.

If the reincorporation of confined population is carried out at a lower rate (30% higher than confinement rate), the best result is obtained. However, it should be emphasized that this progressive reincorporation would be more optimal -in terms of a lower burden of disease-if carried out not at 30 days since confinement but at least at 45 days. This can be seen in the left panel of Figure 4, where a second epidemic peak is not observed during the remaining of 2020. Therefore, it would be possible to bring under control the current outbreak in satisfactory conditions, avoiding the appearance of a second wave of cases of Covid-19 during 2020. The most substantial difference therefore occurs based on the time at which the progressive reincorporation of the confined population is allowed. This results in a reduction from 250, 000 active cases to about 100, 000 active cases, if the start of progressive de-confinement is delayed from April 14 to April 29.

Finally, by way of illustration, Figure 5 shows different dynamics in active and total cases for a generic model scenario in which the ratio *α/τ* varies decreasing from 1 to 0.5. As it can be seen in the upper part of the figure, when the ratio is 1 the effect of prolonging confinement is seen both as an increase in the size of the peak of active cases as well as for the total cases. This behavior is repeated in the other dynamics up to a maximum when the de-confinement rate (*τ*) doubles with respect to the protection rate (*α*). This effect is mainly noticeable in the total number of cases as well as in the appearance and dimension of the second wave of cases. This multipanel figure strongly suggests that releasing a large percentage of the population after the lockdown, results in larger and earlier peaks in the current and next epidemic waves.

**Figure 5.**
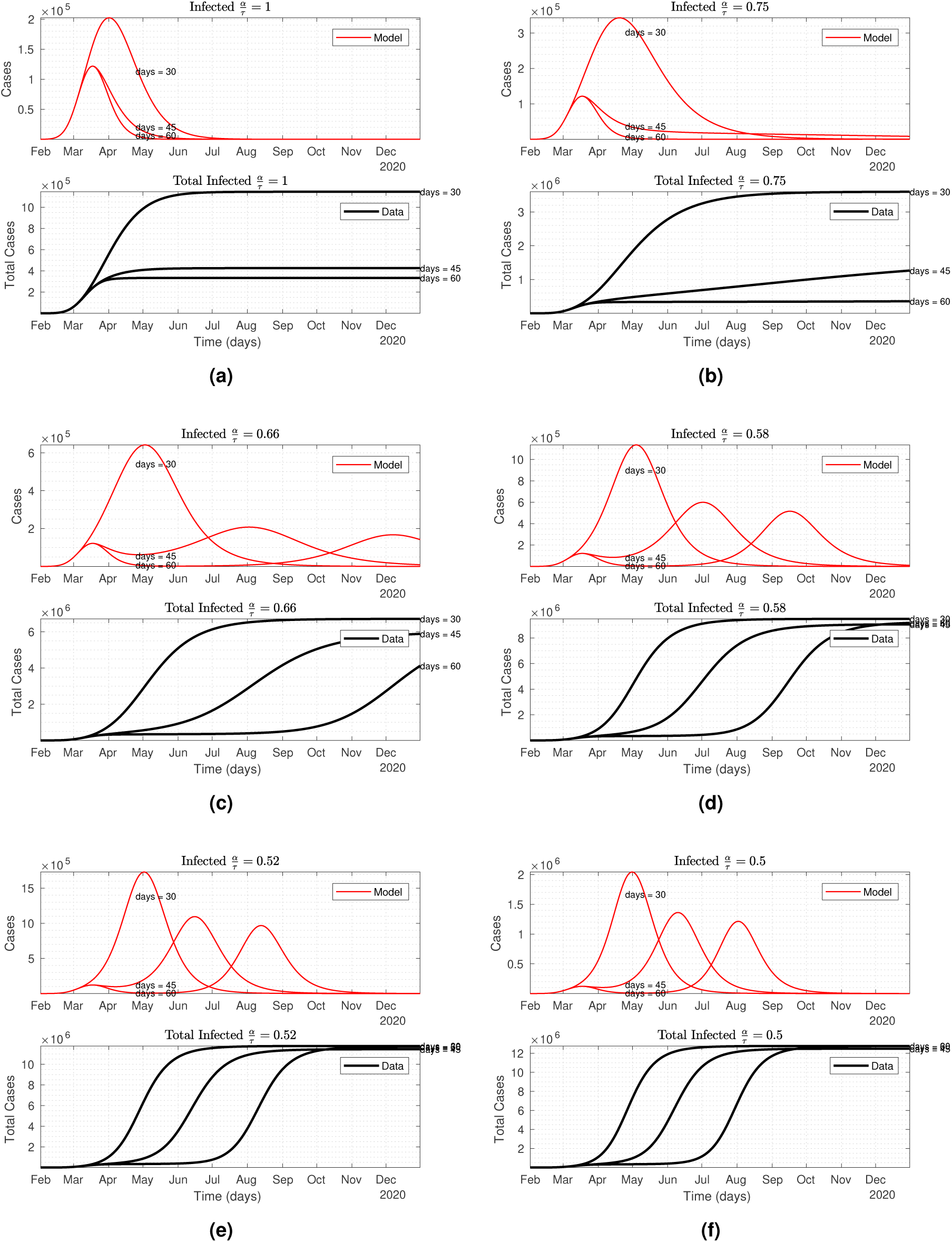
Different ratios for *α/τ* and the effect in the active cases dynamics for a generic case

## Data Availability

All the data included in this manuscript is available in public repositories.

https://www.mscbs.gob.es/profesionales/saludPublica/ccayes/alertasActual/nCov-China/home.htm

## References

1. Worldometers. Worldometers coronavirus (2020).

2. Soper, G. A. The lessons of the pandemic. Science 49, 501–506, DOI: 10.1126/science.49.1274.501 (1919). https://science.sciencemag.org/content/49/1274/501.full.pdf.

3. Martini, M., Gazzaniga, V., Bragazzi, N. & Barberis, I. The spanish influenza pandemic: a lesson from history 100 years after 1918. J. Prev. Medicine Hyg. 60, E64 (2019).

4. Li, R. et al. Substantial undocumented infection facilitates the rapid dissemination of novel coronavirus (sars-cov2).Science (2020).

5. Tapia, M. & Bouza., J. Lo que la pandemia deja al descubierto. el COVID-19 en españa (2020).

6. Zaar, M. H. & Ávila, M.-B. G. El covid-19 en españa y sus primeras consecuencias (2020).

7. Vaguardia, L. Pedro sánchez prepara hoy el inicio de la desescalada del confinamiento en españa (2020).

8. Laurent, L. How do you lift a covid-19 lockdown? ask austria (2020).

9. Organization, W. H. Rolling updates on coronavirus disease (covid-19) (2020).

10. Dropkin, G. COVID-19 UK lockdown forecasts and R0.

11. Peng, L., Yang, W., Zhang, D., Zhuge, C. & Hong, L. Epidemic analysis of covid-19 in china by dynamical modeling.arXiv preprint 2002.06563 (2020).

12. Lopez, L. R. & Rodo, X. A modified seir model to predict the covid-19 outbreak in spain and italy: simulating control scenarios and multi-scale epidemics. medRxiv DOI: 10.1101/2020.03.27.20045005 (2020). https://www.medrxiv.org/content/early/2020/04/12/2020.03.27.20045005.full.pdf.

